# Assessing genetic factors, presenting symptoms, and comorbidities in ovarian cancer diagnosis and survival: a retrospective study

**DOI:** 10.64898/2026.08.11.26360203

**Authors:** Sooah Ko, Maya Demirchian, Edgar Diaz Miranda, Carlye Goldenberg, Kavita Krell, Emma Parry, Mark Hunter, Lisa Brennaman, Amanda Hull, Cassandra Voth, Lei Lei

## Abstract

**Objective:** The purpose of this study is to determine how family history of cancer, genetic mutations, presenting symptoms, and comorbidity burden collectively influence cancer outcomes in patients with epithelial ovarian cancer.

**Methods:** A retrospective analysis was conducted on all patients with epithelial ovarian cancer treated at the University of Missouri and Ellis Fischel Cancer Center between 2008 and 2024. Patient charts were reviewed for histological subtypes, stage of cancer, status of metastasis, CA-125 values, presenting symptoms, comorbidities, family history of cancer, genetic mutations, and survival outcome. Cox regression and association analyses were performed.

**Results:** In this cohort of patients, comorbidities and genetic mutations did not influence ovarian cancer survival. While histological subtypes, CA-125 levels, and cancer stage remained strongly associated with survival. Significant associations were observed between certain presenting symptoms and cancer histological subtype, a family history of breast cancer, stage of cancer at diagnosis, the status of metastasis, and CA-125 levels.

**Conclusion:** Comorbidities and genetic mutations were not significantly associated with ovarian cancer survival. Presenting symptoms were associated with several clinical and pathological variables linked to ovarian cancer diagnosis.

## Introduction

Ovarian cancer remains one of the most lethal gynecologic malignancies worldwide [1]. Globally, more than 300,000 new ovarian cancer cases are diagnosed, and more than 200,000 deaths occur per year [2]. The high mortality associated with ovarian cancer is largely attributable to late-stage diagnosis, the absence of effective screening strategies, and the aggressiveness of high-grade serous carcinoma. Epithelial ovarian cancer (EOC), which accounts for approximately 90% of ovarian malignancies, represents a heterogeneous group of diseases rather than a single entity [3]. These tumors differ substantially in histologic features, molecular alterations, clinical behavior, and treatment response.

Genetic mutations play a critical role in ovarian cancer risk and therapeutic response. Mutations in *BRCA1* and *BRCA2* are present in approximately 15-20% of cases and are associated with defects in homologous recombination double-stranded DNA repair [4]. Thus, these tumors have increased sensitivity to platinum-based chemotherapy and PARP inhibitors. Additional mutations, including TP53 mutations, as well as mutations that impair homologous recombination and dysregulate the PI3K/AKT pathway, further contribute to disease heterogeneity [5]. Molecular characterization has become increasingly important for prognostics, treatment selection, and the development of targeted therapies.

Ovarian cancer has historically been described as a “silent disease”. However, accumulating evidence demonstrates that most patients experience symptoms prior to diagnosis. The challenge lies in the fact that symptoms are nonspecific and common in benign conditions and are therefore frequently attributed to gastrointestinal or urinary etiologies. Several studies have shown that symptom frequency and persistence are important factors in distinguishing ovarian cancer from benign conditions [6]. The ovarian cancer symptom index identified key symptoms, including bloating, pelvic or abdominal pain, early satiety, and urinary frequency or urgency, that were more strongly associated with malignancy [7]. Symptoms occurring more than 12 times per month and present for less than one year are associated with an increased likelihood of ovarian cancer [7].

Comorbidities are prevalent in approximately 35-40% of epithelial ovarian cancer patients and play a role in patient prognosis. The most common comorbidities include hypertension, diabetes, and cardiovascular disease. Comorbidity rates are significantly higher among older patients and can impact both treatment selection and response to treatment [8,9].

The purpose of this study is to determine how genetic factors (including family history of cancer and genetic mutations), presenting symptoms, and comorbidity burden collectively influence cancer outcomes in patients with epithelial ovarian cancer. Understanding how these factors interact may provide insights into earlier recognition of epithelial ovarian cancer, risk stratification, and the development of improved prognostic models.

## Materials and Methods

### 2.1. Patient data collection and categorization

A retrospective chart review was conducted on all patients diagnosed with epithelial ovarian cancer who were treated at the University of Missouri and Ellis Fischel Cancer Center between 2008 and 2024 (IRB#2099034). Patients were identified using the ICD-10 site code “C56.9 - Ovary” from the University of Missouri Health Care Cancer Registry. The initial cohort included 330 cases, which included all patients diagnosed with ovarian cancer who underwent treatment or opted for no treatment at the University of Missouri. Cases were removed if they were non-primary cases, if there was no confirmation of primary epithelial ovarian cancer, if there was a simultaneous diagnosis of multiple primary cancers, or if the initial treatment occurred at another hospital. The final dataset comprised 239 patients, of whom 115 patients had genetic test results (**Supplementary Figure 1**). Data were collected using REDCap, a secure web application for creating and managing online surveys and databases. The following information was gathered from clinical notes: patient medical histories, laboratory results, tumor histologic subtype, clinical stage at diagnosis, and presenting symptoms. The patient’s status (alive, deceased, or unknown) was determined based on the last date of contact recorded in the hospital files. Patients with documented contact within the 2 years prior to the study period were classified as ‘alive’; patients with no contact during this period and without a recorded death date were classified as ‘unknown’; patients with a confirmed death date in the hospital records were classified as ‘deceased’. Patients discharged to hospice care were assumed to have died within the same year of discharge, unless the medical records specified otherwise.

### 2.2. Statistical analysis

Data analysis was conducted in the R environment (version 4.3.3; R Core Team, 2024) using the following packages: survival, survminer, dplyr, tidyr, forcats, glmnet, rstatix, DescTools, gtsummary, broom, nlme and car.

### 2.3. Missing values and imputation

The absence of a CA-125 value was rare (6/239; 2.5%) among the patients. Associations between missing values and observed clinical and pathological variables were evaluated using Fisher’s exact tests, with effect sizes quantified by bias-corrected Cramér’s V. Missing values were imputed using a generalized least squares (GLS) regression model. The model was fitted on patients with observed CA-125 values, using histological type, metastasis status, and clinical symptoms/comorbidities as predictors.

### 2.4. Study outcome and censoring

Survival time was defined as the interval between baseline assessment and the occurrence of the event of interest. Patients without an event during follow-up were right-censored at the date of last contact.

### 2.5. Comorbidity assessment

A predefined set of comorbid conditions was evaluated, including hypertension, type 2 diabetes mellitus, cardiovascular disease, gastrointestinal disease, pulmonary disease, renal disease, arthritis, and hyperlipidemia. Each comorbidity was recorded as a binary numerical indicator reflecting presence or absence.

### 2.6. Survival analyses

Univariate Cox proportional hazards (CPH) models were first fitted to evaluate the association between each comorbidity and the hazard of experiencing the event. Hazard ratios and corresponding confidence intervals were estimated for each model. A multivariable Cox model that simultaneously included all comorbidities was then fitted to assess their independent effects. In parallel, a base clinical model was specified, incorporating key clinical and pathological covariates considered essential a priori, namely stage at diagnosis, CA-125 classification, and histological subtype. A full model combining comorbidities with the clinical backbone was subsequently estimated to evaluate the joint contribution of all variables. Model assumptions were systematically evaluated. The proportional hazards assumption was assessed using tests based on scaled Schoenfeld residuals. Multicollinearity among predictors in multivariable models was examined through generalized variance inflation factors (GVIFs).

In individuals (n = 115) with available genetic testing results, survival analyses were repeated to assess the prognostic relevance of mutation in: (1) any of the tested cancer-associated genes (*BRCA1, BRCA2, TP53, PTEN, KRAS, ATM, RAD51C, RAD51D, MLH1, MSH2, MSH6, PMS2, EPCAM*, and *STK11*); and (2) DNA damage repair (DDR)-associated genes (*BRCA1, BRCA2, ATM, RAD51C*, and *RAD51D*). Univariate Cox models were first fitted for both genetic variables. Base clinical models identical to those used in the full cohort were then extended by adding genetic predictors. Model diagnostics, including proportional hazards testing and collinearity assessment, were conducted for all genetic models. Model selection and penalized regression procedures analogous to those applied in the comorbidity analyses were used to identify final multivariable models for genetic predictors. All statistical tests were two-sided, and significance was evaluated at a nominal alpha level of 0.05. Where applicable, resampling-based procedures and cross-validation were used to ensure robustness of inference.

### 2.7. Model selection and penalized regression

To explore alternative model specifications and identify parsimonious models, an information-theoretic approach was applied. Automated model selection was conducted by fitting all permissible sub-models derived from the full model, subject to structural constraints, and ranking them according to Akaike’s Information Criterion. Model averaging was then performed across the candidate set, yielding averaged coefficient estimates and variable importance metrics. In parallel, penalized Cox regression using the least absolute shrinkage and selection operator (LASSO) was implemented as an alternative variable selection strategy. Predictor matrices were constructed using dummy coding for categorical variables, and cross-validation was applied to identify the optimal regularization parameter. Variables with nonzero coefficients at the optimal penalty level were retained as selected predictors. Based on the convergence of results from classical model selection and penalized regression, a final Cox proportional hazards model was specified. This model included clinically relevant covariates and variables consistently identified as influential. Proportional hazards assumptions were re-evaluated for the final model.

### 2.89. Association analyses

Associations between categorical variables were examined using contingency table analyses. Family history of cancer, genetic mutations, and clinical cancer outcomes, including symptoms, histological subtypes, stage at diagnosis, metastasis status, and CA-125 classification, were evaluated pairwise. For each comparison, contingency tables were constructed and assessed for structural validity. Fisher’s exact test with Monte Carlo simulation (1,000,000 replicates) was applied to obtain p-values. For multi-level categorical predictors, pairwise comparisons between levels were conducted when overall associations were identified. Fisher’s exact tests were applied to each pairwise contrast, and p-values were adjusted for multiple testing using the Holm correction to control the family-wise error rate. Separate association analyses were conducted for the full cohort (n = 239) and for the subset of individuals who underwent genetic testing (n = 115). In the subgroup with genetic testing results, mutation (Yes/No) in tested cancer-associated genes and DDR-associated genes were evaluated in relation to cancer clinical outcomes using the same analytical framework.

## Results

### Summary of the patients

A total of 239 patients were categorized based on the stage of cancer at diagnosis, CA-125 level, metastasis status, survival status, cancer histological subtype, genetic mutations, family history of cancer, presenting symptoms, and comorbidities (**Table 1**). CA-125 levels were categorized into groups 1 (under 35 U/ml), 2 (36-499 U/ml), and 3 (over 500 U/ml). Among the study cohort, 62.8% of patients were diagnosed at an advanced stage (stages III and IV); 44.4% had CA-125 levels in the intermediate range (36-499 U/ml); and 53.6% had metastatic disease. The most common histological subtype was serous cystadenocarcinoma, present in 53.1% of patients. Among patients with genetic testing results, 33.0% tested positive for mutations in cancer-related genes, and 75.7% of patients reported a family history of any type of cancer. Among the top ten symptoms examined, pain, including pelvic, abdominal, flank, and back pain, was the most common, experienced by 75.7% of patients. Among the eight comorbidities evaluated, hypertension was the most prevalent, affecting 36.0% of the patients.

**Table 1.** Characteristics of the study cohort (n=239 patients).

| **Variables** | **Classifications** | **No. of patients (%)** |
| --- | --- | --- |
| **Stage at diagnosis** | Advanced (stages III and IV) | 150 (62.8%) |
|  | Early (stages I and II) | 89 (37.2%) |
| **CA-125 classification** | 1 ($\leq 35$ U/ml) | 43 (18.0%) |
|  | 2 (36–499 U/ml) | 106 (44.4%) |
| | 3 ($\geq 500$ U/ml) | 90 (37.7%) |
| **Metastasis** | Yes | 128 (53.6%) |
|  | No | 94 (39.3%) |
|  | Unknown | 17 (7.1%) |
| **Survival status** | Alive | 121 (50.6%) |
|  | Dead | 86 (36.0%) |
|  | Unknown | 32 (13.4%) |
| **Histological subtype of cancer** | Clear cell cystadenocarcinoma (CCC) | 20 (8.4%) |
|  | Unspecified cystadenocarcinoma (UC) | 19 (7.9%) |
|  | Endometrioid adenocarcinoma (EA) | 30 (12.6%) |
|  | Mixed cell adenocarcinoma (MCA) | 18 (7.5%) |
|  | Mucinous cystadenocarcinoma (MC) | 25 (10.5%) |
|  | Serous cystadenocarcinoma (SC) | 127 (53.1%) |
| **Genetic mutations** | Yes (Cancer-associated genes: *BRCA1*, *BRCA2*, *P53*, *PTEN*, *KRAS*, *ATM*, *RAD51C*, *RAD51D*, *MLH1*, *MSH2*, *MSH6*, *PSM2*, *EPCAM*, *STK11*) | 38 (33.0%) |
|  | Yes (DDR*-associated genes: *BRCA1*, *BRCA2*, *ATM*, *RAD51C*, *RAD51D*) | 28 (24.3%) |
| **Family history of cancer** | Yes (any cancer) | 181 (75.7%) |
|  | Yes (ovarian cancer) | 29 (12.1%) |
|  | Yes (breast cancer) | 76 (31.8%) |
| **Symptoms** | 1. Pain (pelvic, abdominal, flank, back) | 157 (65.7%) |
|  | 2. Abdominal bloating | 100 (41.8%) |
|  | 3. Systemic symptoms (weight loss, loss of appetite, tiredness) | 75 (31.4%) |
|  | 4. Nausea/vomiting | 67 (28.0%) |
|  | 5. Change in bowel movements (constipation, diarrhea) | 59 (24.7%) |
|  | 6. Mass felt | 32 (13.4%) |
|  | 7. Gynecological symptoms (change in menses, spotting, vaginal bleeding, discharge, pain with sex) | 31 (13.0%) |
|  | 8. Urinary symptoms (urgency, frequency) | 28 (11.7%) |
|  | 9. Chest symptoms (chest pain, short of breath) | 25 (10.5%) |
|  | 10. GERD** symptoms (indigestion, heart burn) | 6 (2.51%) |
| **Comorbidities** | 1. Hypertension | 86 (36.0%) |
|  | 2. Hyperlipidemia | 51 (21.3%) |
|  | 3. Cardiovascular disease | 39 (16.3%) |
|  | 4. Type 2 diabetes mellitus | 38 (15.9%) |
|  | 5. Gastrointestinal disease | 33 (13.8%) |
|  | 6. Pulmonary disease | 27 (11.3%) |
|  | 7. Arthritis | 22 (9.2%) |
|  | 8. Renal disease | 11 (4.6%) |

\*DDR: DNA damage repair.
\*\*GERD: Gastroesophageal reflux disease symptoms.

### Comorbidities did not significantly affect ovarian cancer survival

In univariate analyses, no comorbidity was significantly associated with survival (**Supplementary Table 1**). Cardiovascular disease showed a weak, non-significant trend toward increased hazard, while all other comorbidities demonstrated hazard ratios close to unity and no significant association. In the multivariable Cox model including only comorbidities, none of the variables were significantly associated with survival, and the overall model fit was poor (concordance index = 0.565). However, in the fully adjusted model including all comorbidities and cancer clinical outcome variables (**Supplementary Table 2**), the covariates tumor stage, CA-125 classification, and histological subtype remained strongly associated with survival, while all comorbidities remained non-significant. Model discrimination was similar to that of the clinical backbone model (concordance index = 0.744). Variance inflation diagnostics indicated no evidence of problematic multicollinearity among predictors, with all scaled GVIF values close to unity and well below commonly used thresholds (GVIF < 5; **Supplementary Table 3**). All permissible sub-models derived from the full model (2,047 in total) were fitted and ranked according to AIC; the top 10 sub-models are listed in **Supplementary Table 4**. Core clinical covariates (cancer stage, CA-125 classification, and histological subtype) were consistently retained across top-ranked models, whereas individual comorbidities showed weaker and less stable support.

Model-averaged CPH analysis was performed to evaluate the independent association between comorbidities and survival while accounting for model uncertainty and adjusting for core clinical covariates. As shown in **Table 2**, among the comorbid conditions evaluated, cardiovascular disease showed the strongest and most consistent association with survival, with a moderate increase in hazard (HR = 1.65, 95% CI: 0.95–2.87), indicating a 65% increase in hazard. In addition, cardiovascular disease showed the highest variable importance among comorbidities (sum of weights = 0.54), although this association did not reach conventional statistical significance. The Kaplan-Meier curves showed that patients with cardiovascular disease had a decrease in survival probability after 144 months (**Supplementary Figure 2B**). Other comorbidities demonstrated low and inconsistent support across the models, with summed Akaike weights below 0.45 and hazard ratios close to unity (**Table 2**). Additionally, in penalized Cox regression with LASSO shrinkage, pulmonary disease and type 2 diabetes mellitus did not retain a non-zero coefficient at the optimal penalty parameter, indicating no independent prognostic contribution beyond random variation (**Supplementary Table 5**).

**Table 2.** Model-averaged Cox regression estimates for comorbidities and clinical covariates associated with survival in ovarian cancer patients.

| Variables | Estimate ± SE | p-value | HR (95% CI) | Sum of weights* |
| --- | --- | --- | --- | --- |
| **Stage at diagnosis** |  |  |  |  |
| Early (ref.) | – | – | – | 1.00 |
| Advanced | 1.386 ± 0.370 | <0.001 | 4.00 (1.94–8.26) | |
| **CA-125 classification** |  |  |  |  |
| 1 (ref.) | – | – | – | 0.88 |
| 2 | 1.267 ± 0.492 | 0.010 | 3.55 (1.35–9.31) | |
| 3 | 1.173 ± 0.495 | 0.018 | 3.23 (1.22–8.52) | |
| **Histological subtype\*\*** |  |  |  |  |
| CCC (ref.) | – | – | – | 0.98 |
| UC | 1.650 ± 0.585 | 0.005 | 5.21 (1.65–16.4) | |
| EA | -0.560 ± 0.686 | 0.414 | 0.57 (0.15–2.19) | |
| MCA | 0.988 ± 0.583 | 0.090 | 2.69 (0.86–8.42) | |
| MC | 0.110 ± 0.652 | 0.867 | 1.12 (0.31–4.00) | |
| SC | 0.270 ± 0.498 | 0.588 | 1.31 (0.49–3.48) | |
| **Comorbidities (ref = 'No')** |  |  |  |  |
| Cardiovascular disease | 0.503 ± 0.281 | 0.073 | 1.65 (0.95–2.87) | 0.54 |
| Renal disease | -0.713 ± 0.533 | 0.181 | 0.49 (0.17–1.39) | 0.44 |
| Hyperlipidemia | -0.397 ± 0.303 | 0.190 | 0.67 (0.37–1.22) | 0.40 |
| Arthritis | -0.485 ± 0.479 | 0.312 | 0.62 (0.24–1.57) | 0.32 |
| Gastrointestinal disease | 0.319 ± 0.349 | 0.360 | 1.38 (0.69–2.73) | 0.30 |
| Hypertension | -0.212 ± 0.254 | 0.403 | 0.81 (0.49–1.33) | 0.29 |
| Pulmonary disease | 0.291 ± 0.366 | 0.427 | 1.34 (0.65–2.74) | 0.27 |
| Type 2 diabetes mellitus | -0.184 ± 0.342 | 0.591 | 0.83 (0.43–1.63) | 0.24 |

\*Sum of Akaike's weight of sub-models in which each variable was contained (n = 1,023 sub-models by each variable).
\*\*Histological subtype: clear cell cystadenocarcinoma (CCC), unspecified cystadenocarcinoma (UC), endometrioid adenocarcinoma (EA), mixed cell adenocarcinoma (MCA), mucinous cystadenocarcinoma (MC), serous cystadenocarcinoma (SC).

### Genetic mutations did not significantly affect ovarian cancer survival

In univariate analyses, no genetic mutation variables (cancer-associated genes or DDR-associated genes) were significantly associated with survival (**Supplementary Table 6**). In the fully adjusted models that included genetic mutation and clinical backbone variables (**Supplementary Table 7**), only CA-125 classification remained strongly associated with survival, with model discrimination similar to that of the clinical backbone model (0.768 and 0.775, respectively). Variance inflation diagnostics indicated no evidence of potential multicollinearity among predictors, with all scaled GVIF values close to unity and well below the commonly used thresholds (GVIF < 5; **Supplementary Table 8**).

All allowable sub-models derived from the full model were fitted and ranked according to AIC. The top 5 sub-models per genetic mutation variable are listed in **Supplementary Table 9**. CA-125 classification was consistently retained across top-ranked models, whereas genetic mutation variables showed weaker and less stable support. Model-averaged CPH analysis was performed to evaluate the independent association of each genetic mutation variable with survival, while accounting for model uncertainty and adjusting for cancer clinical covariates. The mutation in any of the 14 tested genes presented low and inconsistent support across the models, with summed Akaike weights of 0.30 and 95% CIs for hazard ratios including the unity (**Table 3**). This indicates that the overall mutation panel is not robustly associated with survival after accounting for clinical covariates and model uncertainty. A marginal association between survival and mutations in DDR-associated genes was observed, with a moderate decrease in hazard (HR = 0.46, 95% CI: 0.19–1.15). In addition, mutations in DDR-associated genes presented greater importance (sum of weights = 0.58) than those in cancer-associated genes (0.30; **Supplementary Table 10**). The Kaplan-Meier curves showed that patients with mutations in DDR-associated genes had an increased probability of survival after 48 months (**Supplementary Figure 2C**). In our analysis, core cancer clinical variables were included in the model to ensure appropriate adjustment. Additionally, in penalized Cox regression with LASSO shrinkage, only tumor stage and CA-125 classification retained a non-zero coefficient at the optimal penalty parameter. This supports the result that mutations in cancer-associated genes have no independent prognostic value beyond random variation, and that mutations in DDR-associated genes have only a marginal contribution (**Supplementary Table 11**).

**Table 3.**
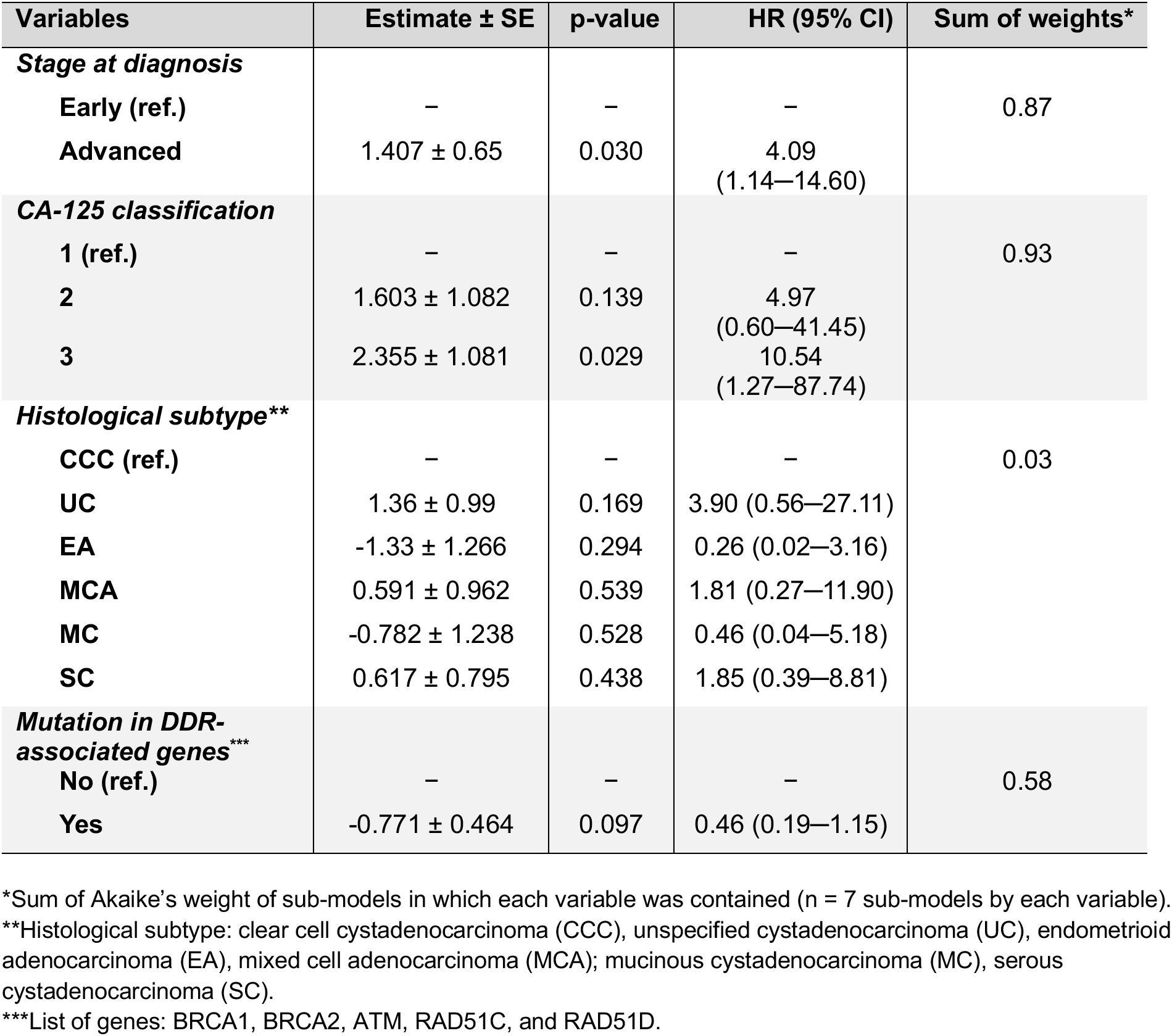
Model-averaged Cox regression estimates for mutations in DDR-associated genes and clinical covariates associated with survival in ovarian cancer patients.

### Associations between genetic factors and ovarian cancer subtypes

Our study cohort included 181 individuals with a family history of any type of cancer, 29 with a family history of ovarian cancer, and 76 with a family history of breast cancer. The analysis showed no significant association between ovarian cancer subtypes and family history of any cancer or ovarian cancer specifically (**Figure 1A, B**). However, we did find a significant association between family history of breast cancer and histological subtypes of ovarian cancer. Notably, a higher percentage of patients with unspecified cystadenocarcinoma (57.9%) had a family history of breast cancer compared to those with serous cystadenocarcinoma (22.6%) **(Figure 1C)**. Additionally, our findings indicated no association between ovarian cancer subtypes and mutations in cancer-associated genes or genes related to DNA damage repair (**Figure 1D, E**)

**Figure 1.**
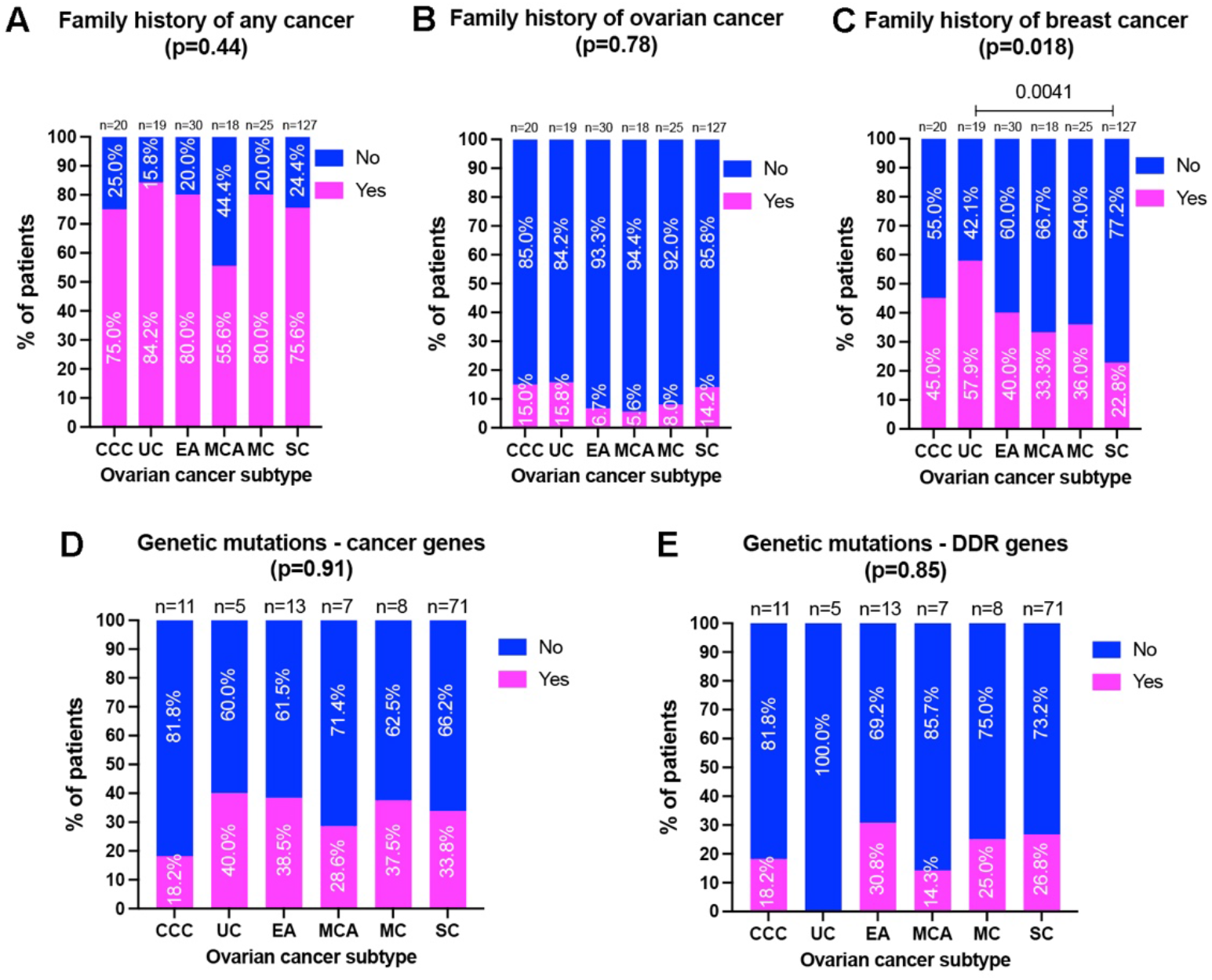
Outcomes of the association analysis between ovarian cancer subtypes and family history of any cancer (A), family history of ovarian cancer (B), family history of breast cancer (C), genetic mutation in cancer-associated genes (D), and genetic mutation in DNA damage repair (DDR)-associated genes.

### Associations between presenting symptoms and cancer clinical outcomes

Our analysis revealed a significant association between cancer subtypes and abdominal bloating or nausea/vomiting. A higher percentage of patients with unspecified cystadenocarcinoma experienced abdominal bloating (73.7%) compared to those with clear cell cystadenocarcinoma (30.0%), endometrioid adenocarcinoma (30.0%), and serous cystadenocarcinoma (34.6%). A higher percentage of patients with mucinous cystadenocarcinoma (76.0%) experienced abdominal bloating compared to patients with endometrioid adenocarcinoma (30.0%) and serous cystadenocarcinoma (34.6%) (**Figure 2A**). Additionally, a lower percentage of patients with clear cell cystadenocarcinoma reported nausea or vomiting (5.0%) than those with unspecified cystadenocarcinoma (52.6%), endometrioid adenocarcinoma (33.3%), mixed cell adenocarcinoma (38.9%), and serous cystadenocarcinoma (28.3%). Furthermore, a higher percentage of patients with unspecified cystadenocarcinoma (52.6%) experienced nausea or vomiting compared to those with mucinous cystadenocarcinoma (12.0%) (**Figure 2B**).

**Figure 2.**
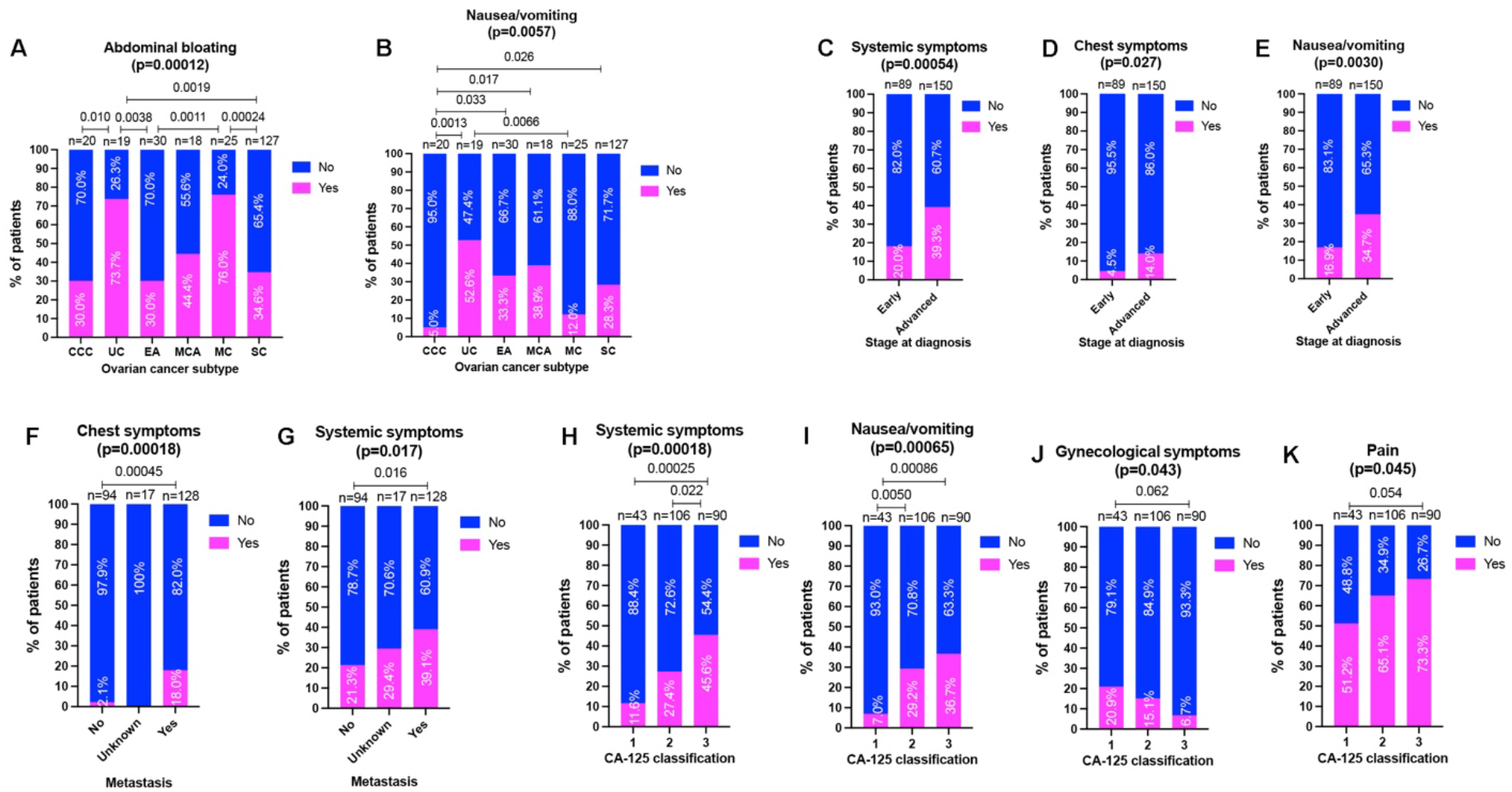
Significant associations were observed between ovarian cancer subtypes and abdominal bloating (A), and nausea/vomiting (B); between stage at diagnosis and systemic symptoms (C), chest symptoms (D), and nausea/vomiting (E); between metastasis and chest symptoms (F), and systemic symptoms (G); and between CA-125 classification and systemic symptoms (H), nausea/vomiting (I), gynecological symptoms, and pain (K).

We observed significant associations between cancer stage at diagnosis and the presenting symptoms. Among patients with advanced-stage ovarian cancer (n=150), a greater percentage experienced systemic symptoms (39.3%), chest symptoms (14.0%), and nausea/vomiting (34.7%). Among patients with early-stage ovarian cancer (n=89), the percentages of patients with the above symptoms were 20%, 4.5%, and 16.9%, respectively (**Figure 2C-E**).

We found that metastatic status was significantly associated with both chest symptoms and systemic symptoms. Among the patients with metastatic ovarian cancer (n = 128), 18.0% experienced chest symptoms, while only 2.1% of patients with non-metastatic ovarian cancer (n = 94) reported chest symptoms (**Figure 2F)**. Additionally, 39.1% of patients with metastatic ovarian cancer experienced systemic symptoms, compared to 21.3% of those with non-metastatic ovarian cancer (**Figure 2G**).

We found that CA-125 classification was significantly associated with systemic symptoms, nausea/vomiting, gynecological symptoms, and pain. Among patients with high CA-125 levels (classification 3), 45.6% experienced systemic symptoms, compared to only 11.6% of patients with low CA-125 levels (classification 1) (**Figure 2H**). 36.7% of patients in the high CA-125 group reported nausea or vomiting, compared with just 7.0% in the low CA-125 group (**Figure 2I**). Only 6.7% of patients with high CA-125 levels reported gynecological symptoms, whereas 20.9% of patients with low CA-125 levels did (**Figure 2J**). Furthermore, 73.3% of patients with high CA-125 levels experienced pain, in contrast to 51.2% of patients with low CA-125 levels (**Figure 2K**).

## Discussion

Survival of EOC appears to be driven more strongly by tumor burden and less by baseline comorbidities. This study contributes to the current literature in support of the use of initial staging and CA-125 levels to stratify EOC mortality risk. While this study did not find a significant difference in survival rates based on patient comorbidities, cardiovascular disease was the comorbidity most strongly associated with survival. Additionally, survival trends for patients with cardiovascular disease were found to be worse compared to those of patients without cardiovascular disease. Previous studies have demonstrated a worse prognosis associated with comorbidities [10]. Patients with significant coexisting conditions often have decreased ability to tolerate aggressive cytoreductive surgery and chemotherapy. Cardiovascular disease was found to be associated with a significant increase in ovarian cancer mortality [11,12]. Various tools have been developed to predict mortality associated with comorbidities. Specifically, the Ovarian Cancer Comorbidity Index predicts survival in ovarian cancer based on age and five conditions: coronary artery disease, hypertension, COPD, diabetes, and dementia [13]. Further clarification on which comorbidities are associated with survival is important to help guide the development of these risk assessment tools. These tools can be used to quickly assess comorbidity burden and guide treatment plans for patients newly diagnosed with EOC.

Symptoms of EOC are vague and appear to occur at varying frequencies across subtypes. Our study revealed that the prevalence of systemic symptoms, chest symptoms, and nausea/vomiting was significantly higher in those with advanced-stage EOC, the presence of metastasis, and higher initial CA-125 values. These findings are consistent with the understanding that EOC symptoms are typically mild and vague until advanced stages of cancer. In contrast, the presence of gynecological symptoms was significantly higher among patients with lower initial CA-125 values. This suggests that increased emphasis might be placed on identifying these symptoms to improve early detection of EOC. Numerous studies have sought to identify the most common presenting symptoms of EOC; however, the challenge remains that the symptoms identified are non-specific and, when considered in isolation, lack sufficient sensitivity and specificity for effective screening. The National Institute for Health and Care Excellence has developed guidelines for referring patients for further workup based on symptom presentation [14]. Future research is needed to more precisely characterize symptom patterns associated with EOC.

In this cohort, no clear relationships were observed between tumor histological subtype and genetic factors. These findings suggest that tumor subtype cannot be readily predicted from family history or mutation status alone. One notable exception was a significant association between family history of breast cancer and ovarian cancer and subtype, which may reflect shared hereditary risk pathways, particularly those related to *BRCA* mutations. We found that mutations in cancer-associated genes or DDR-associated genes did not demonstrate a statistically significant impact on survival, although a trend toward improved survival was observed among patients who tested positive for these mutations. This finding can be substantiated by existing data that tumors with defects in homologous recombination repair can exhibit increased sensitivity to platinum-based chemotherapy and poly ADP-ribose polymerase (PARP) inhibitors [15]. Additionally, patients with hereditary cancer syndromes may undergo earlier surveillance or intervention, which could influence outcomes, consistent with previous findings that improved outcomes were observed in patients with BRCA-mutated ovarian cancer [16].

One strength of this study is that it integrates multiple factors, including clinical cancer outcomes, symptom profiles, patient comorbidities, tumor markers, histologic subtype, and genetic alterations, within a single analysis. Additionally, the use of a real-world dataset enhances the generalizability of our findings to routine practice settings, unlike selected clinical trial populations. The model-averaging approach of our multivariate analysis also strengthens our findings by accounting for model uncertainty. However, our study also has some limitations. Its retrospective nature introduces the potential for selection bias and limits the ability to establish causal relationships. Additionally, the sample size, particularly within certain histologic and genetic subgroups, may have limited statistical power to detect significant associations, especially for mutation-specific effects. Because genetic data were not available for all patients, the mutations analyzed may have obscured histologic subtype-specific effects. Treatment-related variables, including surgical outcomes, chemotherapy regimens, and targeted therapies, were not fully accounted for as well and may have influenced survival outcomes. The data were extracted from clinical documentation and may therefore be subject to recall bias or inconsistent reporting, especially for symptom-related data, which were subjectively reported by patients. Lastly, because this study was conducted within a single institutional cohort, the findings may not be fully generalizable to broader populations. Despite these limitations, this study provides a comprehensive evaluation of multiple factors that influence ovarian cancer prognosis, shedding light on the relative importance of tumor-related factors in determining the survival outcomes among patients with ovarian cancer.

## Data Availability

All data produced in the present study are available upon reasonable request to the authors

## Acknowledgments

None

## Figure legends

**Supplementary Figure 1.**
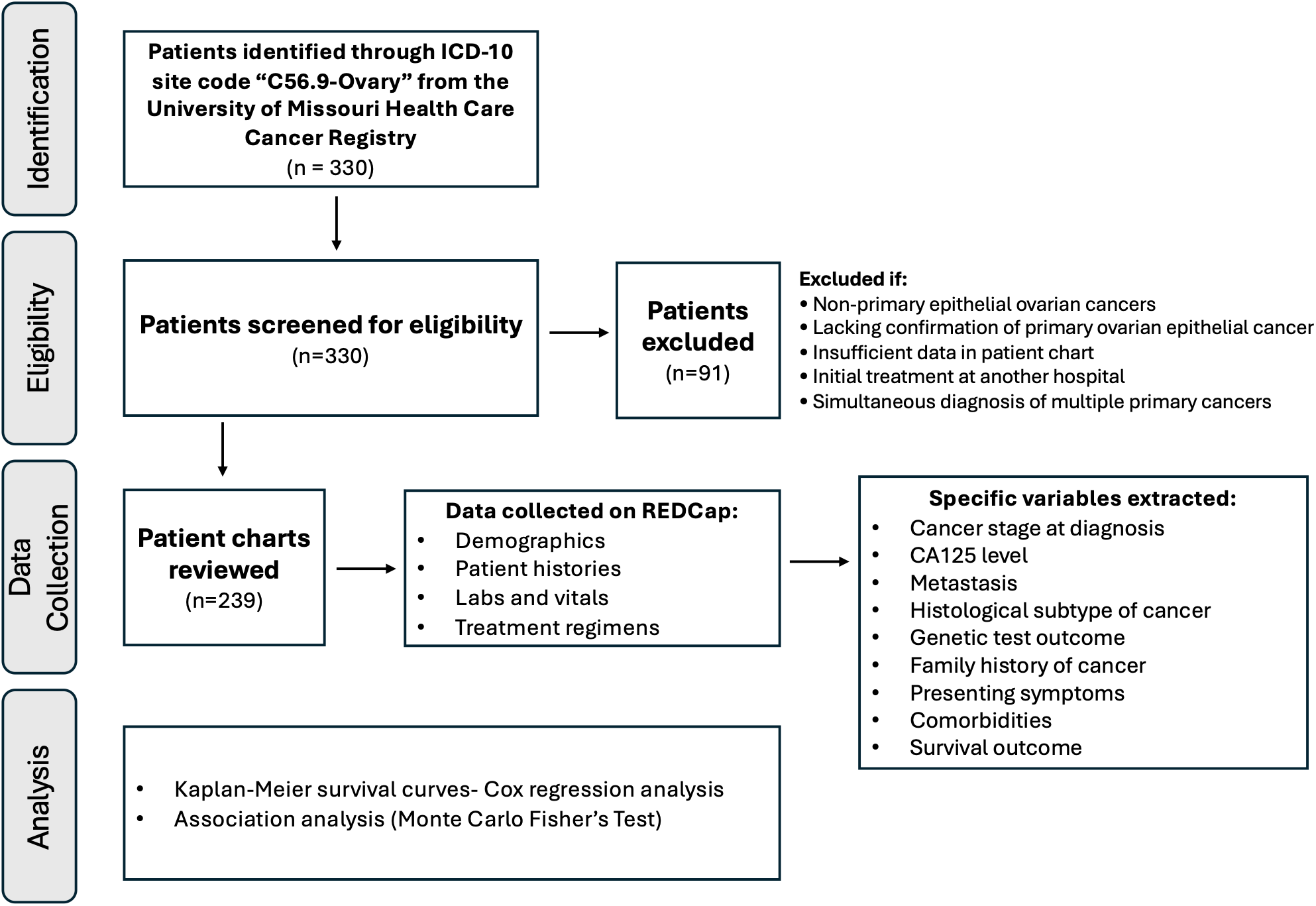
Chart review methodology and patient selection flowchart.

**Supplementary Figure 2.**
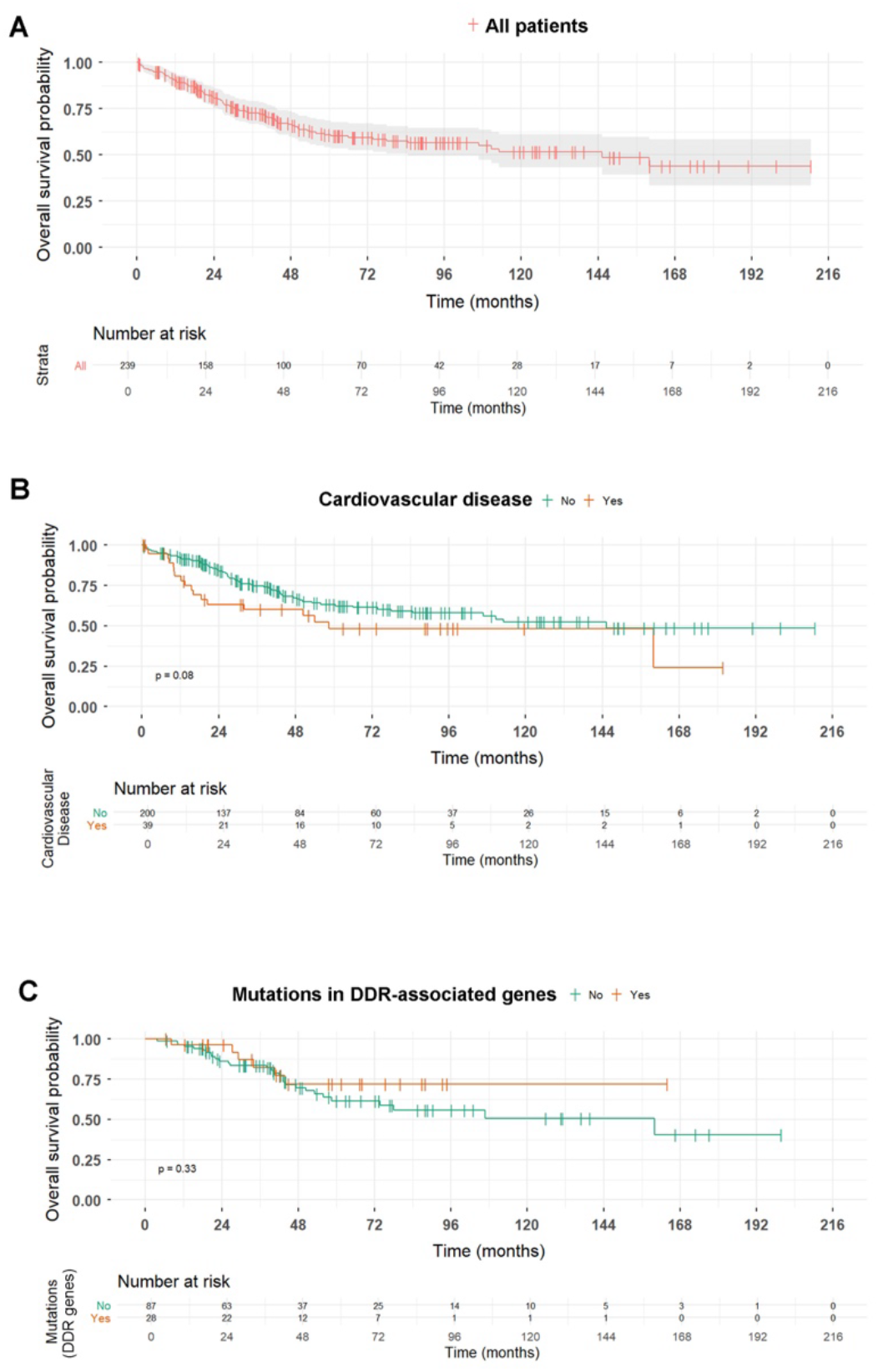
Survival curves over time (in months) based on cardiovascular disease and mutation in DNA damage repair (DDR)-associated genes.

**Supplementary Table 1.** Univariate Cox models for each comorbidity.

| Variables (ref. = 'No') | HR (95% CI) | p-value |
| --- | --- | --- |
| Hypertension | 1.11 (0.72–1.70) | 0.645 |
| Type 2 diabetes mellitus | 1.12 (0.64–1.96) | 0.692 |
| Cardiovascular disease | 1.59 (0.94–2.67) | 0.083 |
| Gastrointestinal disease | 1.30 (0.75–2.23) | 0.351 |
| Pulmonary disease | 1.16 (0.62–2.20) | 0.642 |
| Renal disease | 1.22 (0.50–3.03) | 0.661 |
| Arthritis | 0.81 (0.33–2.01) | 0.651 |
| Hyperlipidemia | 0.81 (0.47–1.39) | 0.435 |

**Supplementary Table 2.** Cox model regression hazard ratios (HR) for comorbidities and clinical covariates associated with survival in ovarian cancer.

| Variables | Base Model<br>(HR [95% CI]; p) | Comorbidity-only<br>Model (HR [95% CI]; p) | Full Model<br>(HR [95% CI]; p) |
| --- | --- | --- | --- |
| **Stage at diagnosis** |  |  |  |
| Early (ref.) | – | – | – |
| Advanced | 3.66 (1.81–7.41); <0.001 | – | 4.14 (2.03–8.44); <0.001 |
| **CA-125 classification** |  |  |  |
| 1 (ref.) |  | – |  |
| 2 | 3.29 (1.27–8.56); 0.014 | – | 3.91 (1.47–10.36); 0.006 |
| 3 | 3.02 (1.15–7.91); 0.025 | – | 3.53 (1.32–9.43); 0.012 |
| **Histological Subtype\*** |  |  |  |
| CCC (ref.) | – | – | – |
| UC | 4.36 (1.44–13.21); 0.009 | – | 5.98 (1.87–19.17); 0.003 |
| EA | 0.55 (0.15–2.09); 0.383 | – | 0.54 (0.14–2.12); 0.374 |
| MCA | 2.46 (0.8–7.57); 0.118 | – | 3.19 (0.99–10.25); 0.051 |
| MC | 1.04 (0.3–3.62); 0.954 | – | 1.19 (0.33–4.38); 0.789 |
| SC | 1.27 (0.48–3.33); 0.630 | – | 1.34 (0.5–3.56); 0.561 |
| **Comorbidities (ref = 'No')** |  |  |  |
| Hypertension | – | 1.15 (0.7–1.88); 0.588 | 0.83 (0.49–1.39); 0.474 |
| Type 2 diabetes mellitus | – | 1.09 (0.59–2.02); 0.784 | 1 (0.5–2); 0.989 |
| Cardiovascular disease | – | 1.5 (0.85–2.67); 0.164 | 1.56 (0.88–2.76); 0.125 |
| Gastrointestinal disease | – | 1.26 (0.67–2.38); 0.474 | 1.55 (0.8–3.02); 0.193 |
| Pulmonary disease | – | 0.95 (0.48–1.88); 0.875 | 1.23 (0.6–2.5); 0.570 |
| Renal disease | – | 1.12 (0.41–3.01); 0.828 | 0.44 (0.15–1.29); 0.134 |
| Arthritis | – | 0.8 (0.31–2.04); 0.636 | 0.64 (0.24–1.67); 0.360 |
| Hyperlipidemia | – | 0.71 (0.38–1.31); 0.273 | 0.72 (0.38–1.34); 0.293 |

\*Histological subtype: clear cell cystadenocarcinoma (CCC); unspecified cystadenocarcinoma (UC); endometrioid adenocarcinoma (EA); mixed cell adenocarcinoma (MCA); mucinous cystadenocarcinoma (MC); serous cystadenocarcinoma (SC).

**Supplementary Table 3.** Generalized variation inflation factor (GVIF) of the full Cox model regression for comorbidities and clinical covariates associated with survival in ovarian cancer.

| Variables | GVIF | DF | Scaled GVIF<br>(GVIF<sup>1/2*DF</sup>) |
| --- | --- | --- | --- |
| Hypertension | 1.4373 | 1 | 1.1989 |
| Type 2 diabetes mellitus | 1.5201 | 1 | 1.2329 |
| Cardiovascular disease | 1.1885 | 1 | 1.0902 |
| Gastrointestinal disease | 1.4767 | 1 | 1.2152 |
| Pulmonary disease | 1.2090 | 1 | 1.0995 |
| Renal disease | 1.3994 | 1 | 1.1829 |
| Arthritis | 1.1366 | 1 | 1.0661 |
| Hyperlipidemia | 1.3090 | 1 | 1.1441 |
| Tumor stage | 1.2347 | 1 | 1.1112 |
| CA-125 classification | 1.3086 | 2 | 1.0696 |
| Histological subtype | 2.1614 | 5 | 1.0801 |

**Supplementary Table 4.** Top 10 full Cox model-derived sub-models.

| Parameters | Top 10 sub-models |  |  |  |  |  |  |  |  |  |
| --- | --- | --- | --- | --- | --- | --- | --- | --- | --- | --- |
|  | 1 | 2 | 3 | 4 | 5 | 6 | 7 | 8 | 9 | 10 |
| Intercept | + | + | + | + | + | + | + | + | + | + |
| Histological subtype | + | + | + | + | + | + | + | + | + | + |
| Stage at diagnosis | + | + | + | + | + | + | + | + | + | + |
| CA-125 classification | + | + | + | + | + | + | + | + | + | + |
| Cardiovascular disease | + | + | + | + | + |  |  |  | + | + |
| Gastrointestinal disease |  |  |  |  |  |  |  |  |  |  |
| Arthritis |  |  |  |  | + |  |  |  |  | + |
| Hyperlipidemia |  | + |  |  |  |  | + |  | + |  |
| Hypertension |  |  |  | + |  |  |  |  |  |  |
| Pulmonary disease |  |  |  |  |  |  |  |  |  |  |
| Renal disease |  |  | + |  |  |  |  | + | + | + |
| Type 2 diabetes mellitus |  |  |  |  |  |  |  |  |  |  |
| Degrees of freedom | 9 | 10 | 10 | 10 | 10 | 8 | 9 | 9 | 11 | 11 |
| AIC* | 793.9 | 794 | 794.4 | 794.7 | 794.8 | 794.9 | 794.9 | 795 | 795.3 | 795.4 |
| Delta** | 0 | 0.12 | 0.48 | 0.73 | 0.93 | 0.97 | 1.03 | 1.04 | 1.34 | 1.45 |
| Weight*** | 0.027 | 0.026 | 0.021 | 0.019 | 0.017 | 0.017 | 0.016 | 0.016 | 0.014 | 0.013 |

\*Akaike's information criterion;
\*\*Difference of the AIC value from the best model;
\*\*\*Akaike's weight, probability that a given model is the best approximating model (among the candidate set), given the data. The "+" sign indicates the presence of the parameter in the model.

**Supplementary Table 5.** Penalized Cox regression with LASSO shrinkage for comorbidities and clinical covariates associated with survival in ovarian cancer.

| Variable | Estimate after LASSO shrinkage |
| --- | --- |
| Stage at diagnosis (Advanced) | 1.3373 |
| CA-125 classification (2) | 0.5293 |
| CA-125 classification (3) | 0.5263 |
| Histological subtype (UC)* | 1.2937 |
| Histological subtype (EA) | -0.4946 |
| Histological subtype (MCA) | 0.6161 |
| Histological subtype (MC) | . |
| Histological subtype (SC) | . |
| Hypertension (Yes) | -0.0022 |
| Type 2 diabetes mellitus (Yes) | . |
| Cardiovascular disease (Yes) | 0.3135 |
| Gastrointestinal disease (Yes) | 0.1669 |
| Pulmonary disease (Yes) | . |
| Renal disease (Yes) | -0.4038 |
| Arthritis (Yes) | -0.2051 |
| Hyperlipidemia (Yes) | -0.2466 |

\*Histological subtype: clear cell cystadenocarcinoma (CCC); unspecified cystadenocarcinoma (UC); endometrioid adenocarcinoma (EA); mixed cell adenocarcinoma (MCA); mucinous cystadenocarcinoma (MC); serous cystadenocarcinoma (SC).

**Supplementary Table 6.** Univariate Cox models for each genetic mutation variable.

| Variables (ref. = 'No') | HR (95% CI) | p-value |
| --- | --- | --- |
| Mutation in cancer-associated genes* | 0.72 (0.34—1.55) | 0.403 |
| Mutation in DDR-associated genes** | 0.65 (0.27—1.57) | 0.338 |

\*Cancer-associated genes: *BRCA1*, *BRCA2*, *P53*, *Pten*, *KRAS*, *ATM*, *RAD51C*, *RAD51D*, *MLH1*, *MSH2*, *MSH6*, *PSM2*, *EPCAM*, and *STK11*.
\*\*DDR-associated genes: *BRCA1*, *BRCA2*, *ATM*, *RAD51C*, and *RAD51D*.

**Supplementary Table 7.** Cox model regression hazard ratios (HR) for genetic mutations and clinical covariates associated with survival in ovarian cancer.

| Variables | Base Model (HR [95% CI]; p) | Mutation Full Model (HR [95% CI]; p) | HR/DR Full Model (HR [95% CI]; p) |
| --- | --- | --- | --- |
| **Stage at diagnosis** |  |  |  |
| Early (ref.) | – | – | – |
| Advanced | 2.70 (0.75–9.77); 0.130 | 2.65 (0.73–9.61); 0.138 | 2.65 (0.73–9.64); 0.139 |
| **CA-125 classification** |  |  |  |
| 1 (ref.) | – | – | – |
| 2 | 6.50 (0.78–54.25); 0.084 | 6.51 (0.78–54.13); 0.083 | 7.09 (0.85–59.45); 0.071 |
| 3 | 8.52 (1.04–69.48); 0.046 | 8.66 (1.06–70.78); 0.044 | 10.04 (1.21–83.51); 0.033 |
| **Histological subtype\*** |  |  |  |
| CCC (ref.) | – | – | – |
| UC | 3.20 (0.47–21.76); 0.234 | 3.07 (0.45–21.23); 0.252 | 2.54 (0.36–17.78); 0.347 |
| EA | 0.21 (0.02–3.00); 0.281 | 0.27 (0.02–3.05); 0.287 | 0.27 (0.02–3.07); 0.290 |
| MCA | 1.67 (0.27–10.33); 0.583 | 1.60 (0.25–10.05); 0.619 | 1.41 (0.22–9.02); 0.719 |
| MC | 0.41 (0.04–4.67); 0.476 | 0.42 (0.04–4.69); 0.478 | 0.40 (0.04–4.48); 0.455 |
| SC | 1.60 (0.34–7.65); 0.555 | 1.58 (0.33–7.53); 0.569 | 1.47 (0.31–7.02); 0.632 |
| **Genetic mutation (ref = 'No')** |  |  |  |
| Cancer-associated genes** | – | 0.83 (0.38–1.81); 0.640 | – |
| DDR-associated genes*** | – | – | 0.60 (0.23–1.44); 0.241 |

\*Histological subtype: clear cell cystadenocarcinoma (CCC); unspecified cystadenocarcinoma (UC); endometrioid adenocarcinoma (EA); mixed cell adenocarcinoma (MCA); mucinous cystadenocarcinoma (MC); serous cystadenocarcinoma (SC).
\*\*Cancer-associated genes: *BRCA1*, *BRCA2*, *P53*, *Pten*, *KRAS*, *ATM*, *RAD51C*, *RAD51D*, *MLH1*, *MSH2*, *MSH6*, *PSM2*, *EPCAM*, and *STK11*.
\*\*\*DDR-associated genes: *BRCA1*, *BRCA2*, *ATM*, *RAD51C*, and *RAD51D*.

**Supplementary Table 8.**
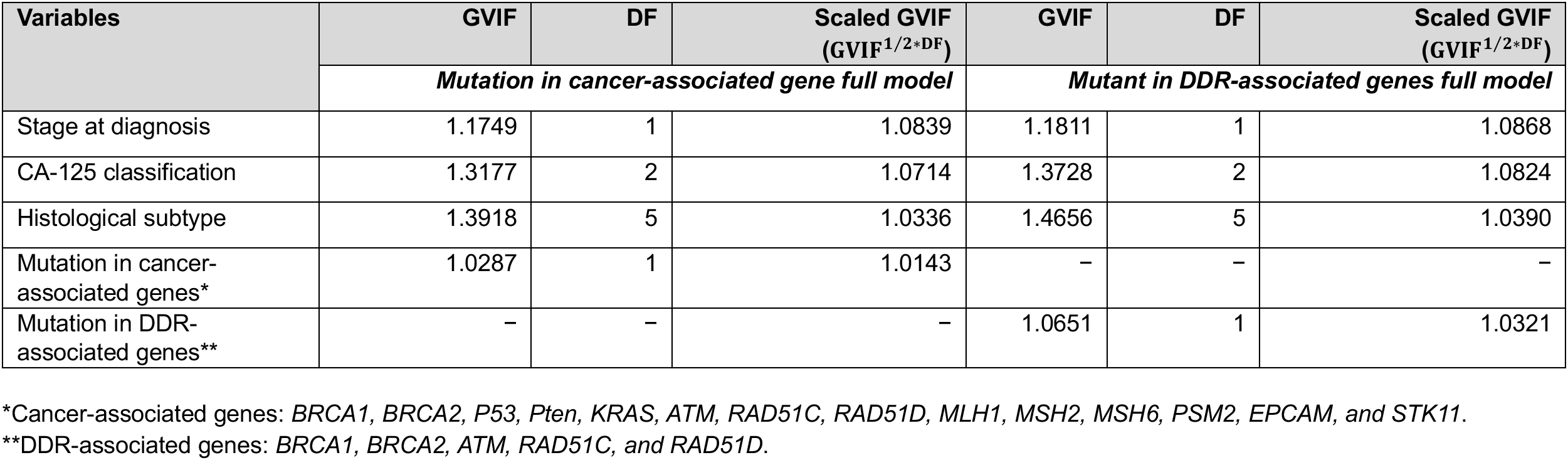
Generalized variation inflation factor (GVIF) of the full Cox model for genetic test variables.

| Variables | GVIF | DF | Scaled GVIF<br>(GVIF<sup>1/2*DF</sup>) | GVIF | DF | Scaled GVIF<br>(GVIF<sup>1/2*DF</sup>) |
| --- | --- | --- | --- | --- | --- | --- |
|  | *Mutation in cancer-associated gene full model* |  |  | *Mutant in DDR-associated genes full model* |  |  |
| Stage at diagnosis | 1.1749 | 1 | 1.0839 | 1.1811 | 1 | 1.0868 |
| CA-125 classification | 1.3177 | 2 | 1.0714 | 1.3728 | 2 | 1.0824 |
| Histological subtype | 1.3918 | 5 | 1.0336 | 1.4656 | 5 | 1.0390 |
| Mutation in cancer-associated genes* | 1.0287 | 1 | 1.0143 | – | – | – |
| Mutation in DDR-associated genes** | – | – | – | 1.0651 | 1 | 1.0321 |

\*Cancer-associated genes: *BRCA1*, *BRCA2*, *P53*, *Pten*, *KRAS*, *ATM*, *RAD51C*, *RAD51D*, *MLH1*, *MSH2*, *MSH6*, *PSM2*, *EPCAM*, and *STK11*.
\*\*DDR-associated genes: *BRCA1*, *BRCA2*, *ATM*, *RAD51C*, and *RAD51D*.

**Supplementary Table 9.** Top 5 full Cox model-derived sub-models for genetic mutation variables.

| Parameters | Mutation in cancer-associated genes<br>Top 5 sub-models |  |  |  |  | Mutation in DDR-associated genes<br>Top 5 sub-models |  |  |  |  |
| --- | --- | --- | --- | --- | --- | --- | --- | --- | --- | --- |
|  | 1 | 2 | 3 | 4 | 5 | 1 | 2 | 3 | 4 | 5 |
| Intercept | + | + | + | + | + | + | + | + | + | + |
| CA 125 classification | + | + |  | + | + | + | + | + |  | + |
| Histological subtype |  |  |  |  |  |  |  |  |  |  |
| Mutation in cancer-associated genes<sup>†</sup> |  | + |  |  | + | n.a. | n.a. | n.a. | n.a. | n.a. |
| Mutation in DDR-associated genes<sup>††</sup> | n.a. | n.a. | n.a. | n.a. | n.a. | + |  | + |  |  |
| Stage at diagnosis | + | + | + |  |  | + | + |  | + |  |
| Degrees of freedom | 3 | 4 | 1 | 2 | 3 | 4 | 3 | 3 | 1 | 2 |
| AIC* | 268.9 | 270.6 | 273.1 | 274 | 274.3 | 268.3 | 268.9 | 271.6 | 273.1 | 274 |
| Delta** | 0 | 1.68 | 4.18 | 5.01 | 5.35 | 0 | 0.70 | 3.38 | 4.87 | 5.71 |
| Weight*** | 0.541 | 0.234 | 0.067 | 0.044 | 0.037 | 0.463 | 0.326 | 0.086 | 0.040 | 0.027 |

\*Akaike's information criterion.
\*\*Difference of the AIC value from the best model.
\*\*\*Akaike's weight, probability that a given model is the best approximating model (among the candidate set), given the data. The “+” sign indicates the presence of the parameter in the model.
<sup>†</sup>Cancer-associated genes: *BRCA1*, *BRCA2*, *P53*, *Pten*, *KRAS*, *ATM*, *RAD51C*, *RAD51D*, *MLH1*, *MSH2*, *MSH6*, *PSM2*, *EPCAM*, and *STK11*.
<sup>††</sup>DDR-associated genes: *BRCA1*, *BRCA2*, *ATM*, *RAD51C*, and *RAD51D*.

**Supplementary Table 10.** Model-averaged Cox regression estimates for any mutation and clinical covariates associated with survival in ovarian cancer.

| Variables | Estimate ± SE | p-value | HR (95% CI) | Sum of weights* |
| --- | --- | --- | --- | --- |
| **Stage at diagnosis** |  |  |  |  |
| Early (ref.) | – | – | – | 0.89 |
| Advanced | 1.457 ± 0.651 | 0.025 | 4.29 (1.20–15.38) |  |
| **CA-125 classification** |  |  |  |  |
| 1 (ref.) | – | – | – | 0.89 |
| 2 | 1.531 ± 1.081 | 0.157 | 4.62 (0.56–38.48) |  |
| 3 | 2.222 ± 1.067 | 0.037 | 9.23 (1.14–74.78) |  |
| **Histological subtype\*\*** |  |  |  |  |
| CCC(ref.) | – | – | – | 0.05 |
| UC | 1.394 ± 0.987 | 0.158 | 4.03 (0.58–27.88) |  |
| EA | -1.323 ± 1.267 | 0.297 | 0.27 (0.02–3.19) |  |
| MCA | 0.625 ± 0.954 | 0.512 | 1.87 (0.29–12.13) |  |
| MC | -0.772 ± 1.238 | 0.533 | 0.46 (0.04–5.24) |  |
| SC | 0.627 ± 0.796 | 0.431 | 1.87 (0.39–8.91) |  |
| **Mutation in cancer-associated genes\*\*\*** |  |  |  |  |
| No (ref.) | – | – | – | 0.30 |
| Yes | -0.365 ± 0.402 | 0.365 | 0.69 (0.32–1.53) |  |

\*Sum of Akaike's weight of sub-models in which each variable was contained (n = 7 sub-models by each variable).
\*\*Histological subtype: clear cell cystadenocarcinoma (CCC); unspecified cystadenocarcinoma (UC); endometrioid adenocarcinoma (EA); mixed cell adenocarcinoma (MCA); mucinous cystadenocarcinoma (MC); serous cystadenocarcinoma (SC).
\*\*\*Cancer-associated genes: *BRCA1*, *BRCA2*, *P53*, *Pten*, *KRAS*, *ATM*, *RAD51C*, *RAD51D*, *MLH1*, *MSH2*, *MSH6*, *PSM2*, *EPCAM*, and *STK11*.

**Supplementary Table 11.**
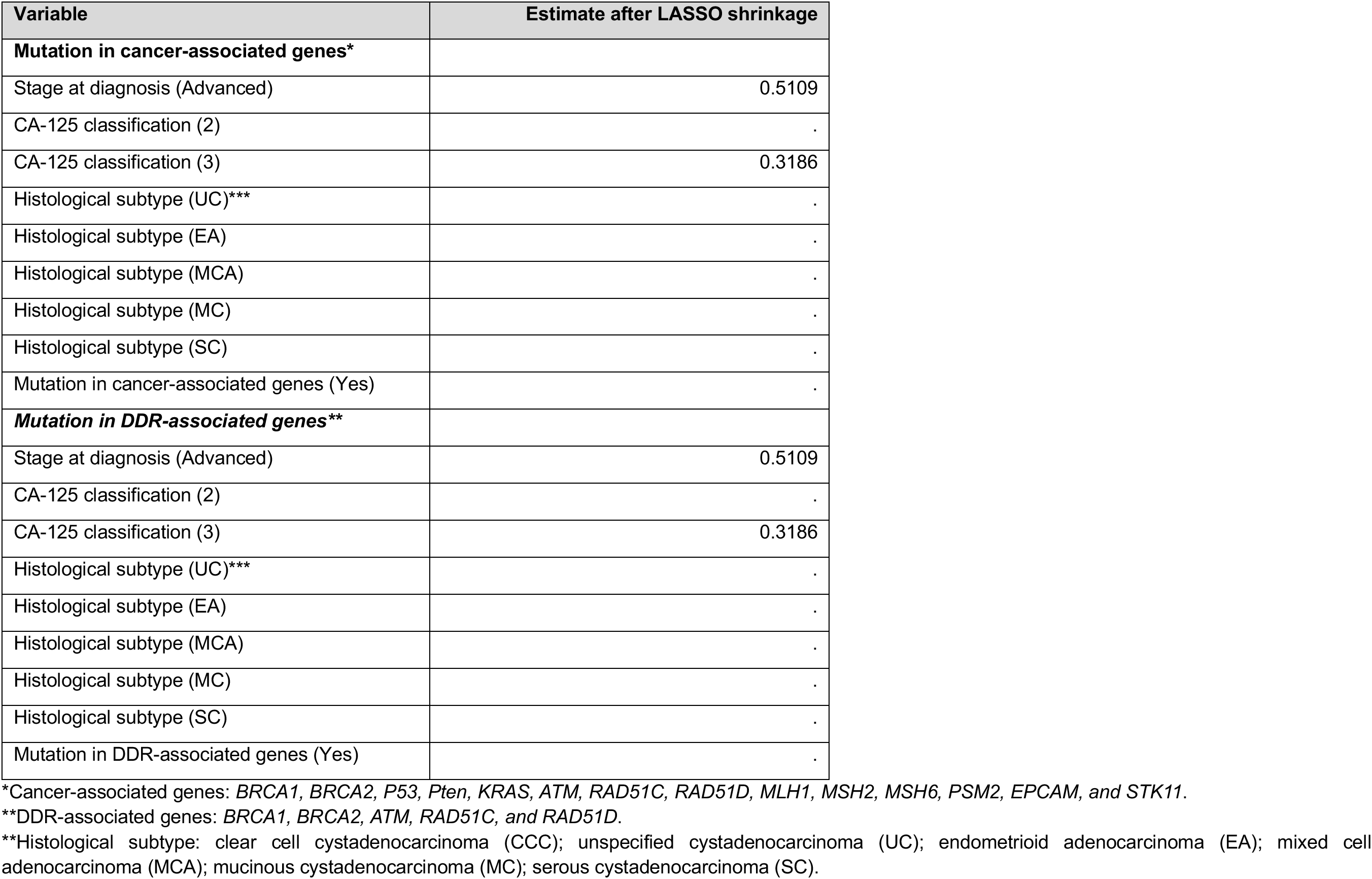
Penalized Cox regression with LASSO shrinkage genetic teste variables and clinical covariates associated with survival in ovarian cancer.

